# Detection of *M. tuberculosis* DNA in TB contacts’ PBMC does not associate with blood RNA signatures for incipient tuberculosis

**DOI:** 10.1101/2023.09.26.23296131

**Authors:** Joshua Rosenheim, Markos Abebe, Mulugeta Belay, Begna Tulu, Dawit Tayachew, Metasebia Tegegn, Sidra Younis, David A. Jolliffe, Abraham Aseffa, Gobena Ameni, Stephen T Reece, Mahdad Noursadeghi, Adrian R Martineau

## Abstract

**Background:** We have previously reported detection of genomic DNA of *Mycobacterium tuberculosis* (Mtb) in peripheral blood mononuclear cells (PBMC) of asymptomatic adults with recent household exposure to an index case of infectious pulmonary tuberculosis (TB). It is not known whether this phenomenon indicates quiescent Mtb infection or incipient TB disease.

**Methods:** We did a cross-sectional study with a nested prospective component, utilizing blood samples collected from a previously studied cohort of TB contacts recruited in Ethiopia to detect eight different blood RNA signatures for incipient TB. Gene expression analysis was performed on baseline blood samples from 24 HIV-uninfected Mtb DNA-negative individuals, 24 HIV-uninfected Mtb DNA-positive individuals, and 24 HIV-infected Mtb DNA-positive participants. Analysis was also performed on follow-up blood samples of 24 HIV-infected participants following completion of isoniazid preventive therapy (IPT), of whom 17 were classified as ‘responders’ to IPT (Mtb DNA undetectable at follow-up) and 7 were classified as ‘non-responders’ to IPT (Mtb DNA detectable at follow-up).

**Results:** Cross-sectional analysis of baseline gene expression data from HIV-uninfected participants revealed no differences in Z-scores between Mtb DNA-positive vs. Mtb DNA-negative participants for any of the eight gene expression signatures investigated. Prospective analyses comparing expression of the same eight signatures in HIV-infected participants before vs. after administration of IPT showed no change in gene expression Z-scores over the course of treatment, either for ‘responders’ or ‘non-responders’ to treatment.

**Conclusions:** Lack of any association between detection of Mtb DNA in PBMC of asymptomatic TB contacts and presence of gene expression signatures investigated suggests that detection of this bacillary biomarker is more likely to represent quiescent Mtb infection than incipient TB disease.

**Funding:** United Kingdom Medical Research Council (Ref. MR/P024548/1)

**RESEARCH IN CONTEXT:** *Evidence before this study□:* We searched PubMed from database inception to Sept 30, 2023, for studies published in any language evaluating associations between blood RNA signatures for tuberculosis (TB) and biomarkers of *Mycobacterium tuberculosis* (Mtb) infection using comprehensive terms for “tuberculosis infection”, “latent tuberculosis”, “transcriptional” and “biomarker”. Several reports have described whole blood transcriptional signatures associating with positive Interferon-Gamma Release Assay results in individuals without clinical or radiological evidence of active tuberculosis, but we did not find studies investigating transcriptional correlates of microbial biomarkers of Mtb infection.

*Added value of this study□:* To our knowledge, this is the first study to investigate whether detection of Mtb DNA in blood of individuals with asymptomatic tuberculosis infection associates with blood RNA signatures for incipient tuberculosis. Cross-sectional analysis of samples from HIV-uninfected TB contacts in Ethiopia revealed no differences in any of eight whole blood gene expression signatures compared between Mtb DNA-positive vs. Mtb DNA-negative participants. Prospective analyses comparing expression of the same eight signatures in HIV-infected participants before vs. after administration of isoniazid preventive therapy showed no change in gene expression signatures over the course of treatment.

*Implications of all the available evidence□:* Lack of any association between detection of Mtb DNA in the blood of asymptomatic TB contacts vs. presence of gene expression signatures for incipient TB provides the best available evidence for bona fide latent Mtb infection.

## INTRODUCTION

Human exposure to *Mycobacterium tuberculosis* (Mtb) is thought to result in a spectrum of outcomes including bacillary clearance, quiescent Mtb infection, incipient tuberculosis, subclinical tuberculosis and active tuberculosis.^1^ Incipient tuberculosis - defined as a prolonged asymptomatic phase of early disease preceding clinical presentation as active disease^2^ – may be distinguished from quiescent Mtb infection by detection of host gene expression signatures in blood, whose presence associates with increased risk of progression to active TB.^3^ Recently, we reported detection of Mtb DNA in peripheral blood mononuclear cells (PBMC) of asymptomatic TB-exposed adults with normal chest radiographs living in Ethiopia. Administration of isoniazid preventive therapy (IPT) reduced the proportion of HIV-infected individuals in whom this signal was detectable, suggesting that detection of Mtb DNA in PBMC may represent the presence of viable bacteria.^4^ Whether these bacteria provoke a host transcriptional response associated with sub-clinical disease is not known. We tested the hypothesis that detection of Mtb DNA in PBMC was associated with increased expression of host blood RNA signatures of incipient disease, using samples collected from study participants before and after administration of IPT.

## METHODS

### Participants

We conducted a cross-sectional study with a nested prospective component as previously described.^4^ Briefly, three groups of asymptomatic adults at risk of latent tuberculosis infection (household TB contacts, farm workers with occupational exposure to bovine TB and HIV-infected people attending an out-patient clinic) and living in or around Addis Ababa, Ethiopia, were invited to give a blood sample for processing as described below. Principal exclusion criteria were age <18 years; symptoms of active TB; haemoglobin concentration <10 g/dL; and presence of any chest radiograph abnormality consistent with active TB. HIV-infected participants who did not have any contra-indication to IPT were offered a 6-month course of 300 mg isoniazid and pyridoxine 50 mg daily and invited to return to give a second blood sample for the same laboratory tests following treatment completion. The study was approved by the Ethiopian National Research Ethics Review Committee, Addis Ababa, Ethiopia (ref 310/253/2017). Written informed consent was obtained from all participants.

### Detection of Mtb DNA in PBMC

PBMC were isolated by density gradient centrifugation from 100 mL blood drawn into sodium heparin tubes, and CD34-positive and -negative PBMC were separated using CD34 MicroBead Ultra-Pure Kits and MS columns (both from Miltenyi Biotec, Bergisch Gladbach, Germany) according to the manufacturer’s instructions. DNA was extracted from CD34-positive and -negative cell pellets using a lysozyme/proteinase K digestion step followed by a Cetyltrimethylammonium bromide (CTAB) / Chloroform-Isoamyl alcohol protocol.^5^ Digital polymerase chain reaction (dPCR) was performed using 10 μl of extract using the QX200 AutoDG Droplet Digital PCR System (Bio-Rad Laboratories, Hercules CA) to detect DNA of two Mtb complex-specific genes: the multi-copy gene insertion sequence (IS)*6110* and the single-copy gene *rpoB*.^6^ Oligonucleotide information and thermal cycling conditions are reported elsewhere.^4^

### Determination of host gene expression signatures

3 mL blood was collected into Tempus Blood RNA Tubes (Thermo Fisher Scientific) to stabilise total RNA for isolation and downstream gene expression analysis, and processed with Tempus Spin RNA Isolation Kits (Thermo Fisher Scientific) to obtain high-quality total RNA. TURBO DNA-free kits (Thermo Fisher Scientific) and GLOBINclear kits (Thermo Fisher Scientific) were then used to remove residual DNA and globin mRNAs, respectively. For genome-wide mRNA sequencing, complementary DNA libraries were generated using Kappa HyperPrep kits (Roche) and sequenced on the Illumina NextSeq 550 system using NextSeq 500/550 High Output 75 Cycle Kits (Illumina), resulting in a median of 25.8 million (range 12.3 – 30.9) 41 bp paired-end reads per sample.

### Statistical analysis

RNA sequencing data were mapped to Ensembl Human GRCh38 release 104 using Kallisto (v0.46.1). Raw read counts were normalised to transcripts per million values, summed at gene level, and annotated with Ensembl gene ID, gene name, and gene biotype using the Bioconductor packages tximport (v1.20.0) and biomaRt (v.2.48.0) in R. Scores for eight incipient TB signatures were calculated as reported elsewhere.^7-13^ Gene expression Z-scores were calculated by standardising to the mean and standard deviation of data from Mtb DNA negative individuals. This fixed the mean Z score of the control group to zero and standard deviation to 1. Statistical power was estimated using PASS software v2022 (NCIS). Unpaired t-tests were used to compare gene expression Z-scores of HIV-uninfected participants in whom Mtb DNA was detected vs. undetected at baseline. In this analysis, a sample size of n=24 per group achieved 92% power to identify a difference in Z score means of ≥1. Paired t-tests were used to compare gene expression Z-scores of HIV-infected participants before vs. after IPT, with stratification by Mtb PCR responder status. In this analysis, a sample size of n=17 responders achieved 97% power, and a sample size of N=7 non-responders achieved 60% power, to identify a difference in Z score means of ≥1. Between-group differential gene expression was investigated using Deseq2 and false discovery rate (FDR)<0.05.^14^ Spearman’s correlation coefficients were used to explore any relationship between Mtb DNA copy number and host blood RNA signatures of TB as continuous variables.

## RESULTS

A total of 284 participants were enrolled between November 22^nd^, 2017 and January 10^th^, 2019. Valid dPCR data (i.e. droplet counts >10,000 per well) were available for paired CD34-positive and CD34-negative PBMC from 197 participants. Gene expression analysis was performed on baseline whole blood samples from randomly selected subsets of 24/33 (72.7%) HIV-uninfected dPCR-negative participants, 24/89 (30.0%) HIV-uninfected dPCR-positive participants, and 24/67 (35.8%) HIV-infected dPCR-positive participants. Table 1 presents characteristics of participants contributing data to the current analysis: median age was 35.0 years (interquartile range 25.5 to 42.5), and 28 (38.9%) were female. Gene expression analysis was also performed on follow-up blood samples of HIV-infected participants following completion of IPT, of whom 17 (70.8%) were classified as ‘responders’ to IPT (Mtb DNA undetectable at follow-up) and 7 (29.2%) were classified as ‘non-responders’ to IPT (Mtb DNA detectable at follow-up).

**Table 1.** Characteristics of participants contributing gene expression data (n=72)

|  |  | **Overall**<br>**(n=72)** | **HIV-**<br>**uninfected,**<br>**Mtb DNA**<br>**undetected at**<br>**baseline**<br>**(n=24)** | **HIV-**<br>**uninfected,**<br>**Mtb DNA**<br>**detected at**<br>**baseline**<br>**(n=24)** | **HIV-**<br>**infected,**<br>**Mtb DNA**<br>**detected at**<br>**baseline**<br>**(n=24)** |
| --- | --- | --- | --- | --- | --- |
| Median age, years (interquartile range) |  | 35.0 (25.5 to 42.5) | 25.5 (22.0 to 35.0) | 34.0 (25.5 to 39.0) | 38.0 (35.0 to 45.0) |
| Female, n (%) |  | 28 (38.9) | 10 (41.7) | 2 (8.3) | 16 (66.7) |
| Study group, n (%) | Recent household exposure to human index case with smear-positive pulmonary TB, Addis Ababa | 21 (29.2) | 10 (41.7) | 11 (45.8) | 0 (0.0) |
|  | Recent occupational exposure to bovine index case with culture-positive bovine TB, recruited in cattle farms | 27 (37.5) | 14 (58.3) | 13 (54.2) | 0 (0.0) |
|  | Attending out-patient HIV clinic, Addis Ababa | 24 (33.3) | 0 (0.0) | 0 (0.0) | 24 (100.0) |
| BCG scar, n (%) |  | 43 (59.7) | 16 (66.7) | 16 (66.7) | 11 (45.8) |
| Current smoker, n (%) |  | 2 (2.8) | 0 (0.0) | 2 (2.8) | 0 (0.0) |
| QuantiFERON-Plus positive, n (%) |  | 39 (54.2) | 12 (50.0) | 16 (66.7) | 11 (45.8) |

Cross-sectional analysis of baseline gene expression data from HIV-uninfected participants revealed no differences in Z-scores between Mtb DNA-positive vs. Mtb DNA-negative participants for any of the eight signatures investigated (Fig. 1A). Prospective analyses comparing expression of the same 8 signatures in HIV-infected participants before vs. after administration of IPT showed no change in gene expression Z-scores over the course of treatment, either for those in whom Mtb DNA was undetectable at follow-up (‘PCR responders’) or for those in whom Mtb DNA was still detected at follow-up (‘PCR non-responders’) (Fig. 1B).

**Figure 1:**
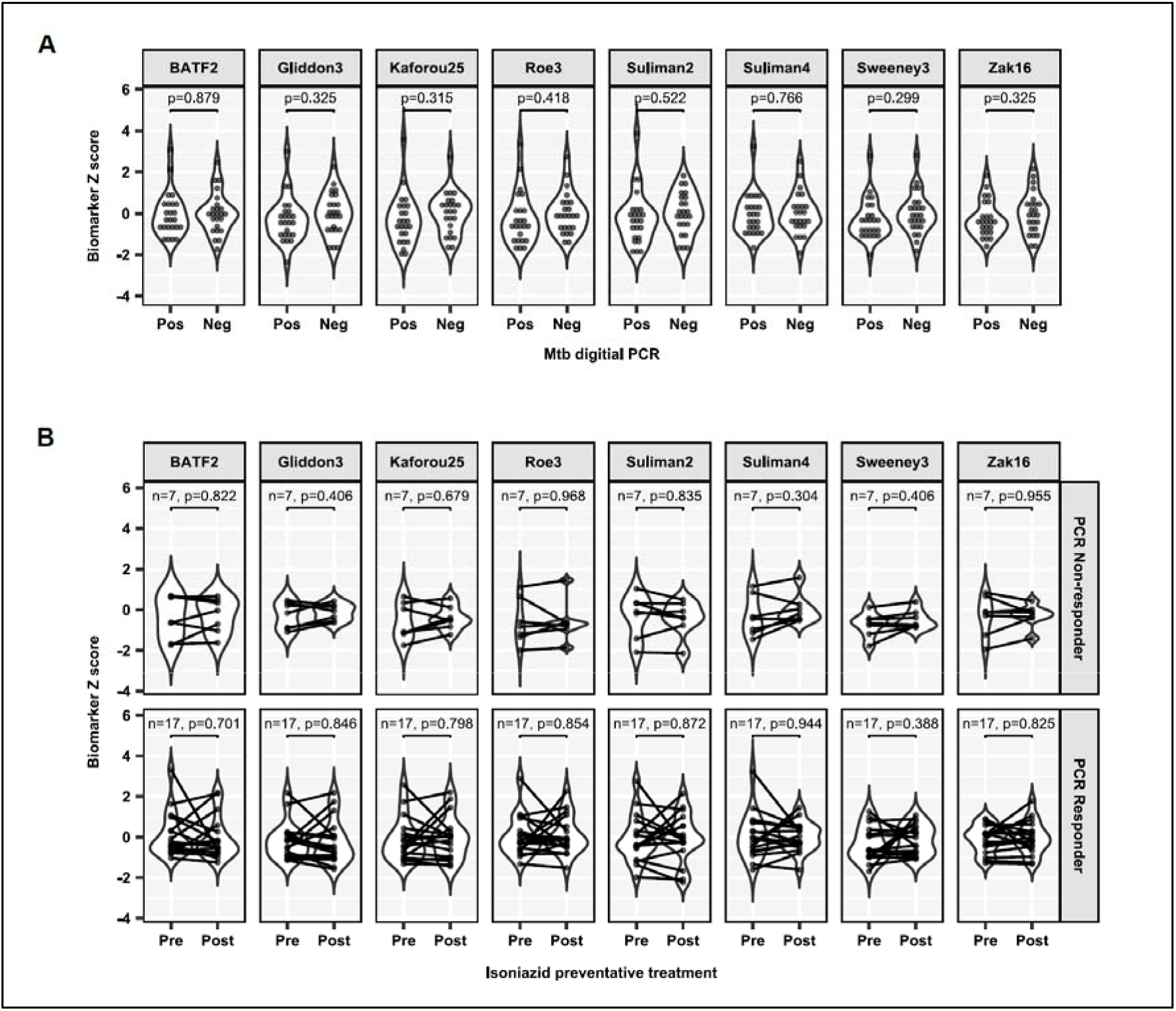
Z-scores for expression of eight different signatures for incipient TB in whole blood of asymptomatic adults in Ethiopia. A, Baseline Z-scores by Mtb DNA status. B, Z-scores before vs. after administration of isoniazid preventive therapy among those in whom Mtb DNA was detected at follow-up (‘PCR non-responders’, top row) and those in whom Mtb DNA was undetectable at follow-up (‘PCR responders’, bottom row).

In a secondary analysis, we explored whether there was any relationship between quantitative levels of Mtb DNA and host blood RNA signatures as continuous variables, across all samples stratified by participant HIV status, blood RNA signatures, PBMC cell subsets and Mtb PCR targets. Correlation coefficients were stronger between pairs of gene signatures and PCR targets within CD34-positive and CD34-negative subsets than between any pairs of RNA signatures and Mtb PCR results (Fig. 2).

**Figure 2:**
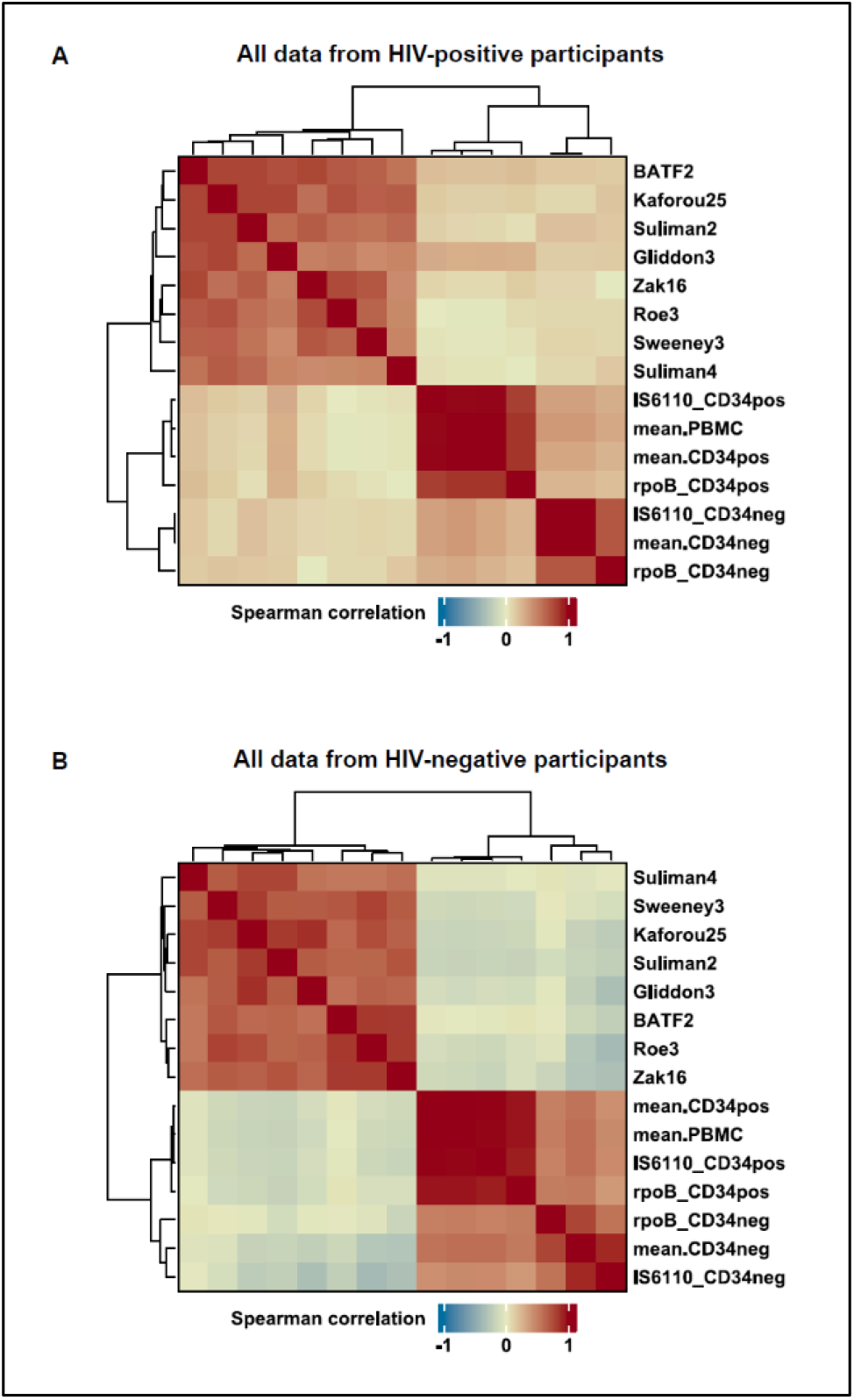
Heatmaps showing correlation between selected blood RNA biomarker measurements and quantitative dPCR measurements. A, HIV-infected participants. B, HIV-uninfected participants.

## DISCUSSION

We present findings of the first study to investigate whether detection of Mtb DNA in peripheral blood of asymptomatic adults at risk of *M. tuberculosis* infection associates with blood RNA signatures for incipient TB. No such associations were found on cross-sectional analysis of baseline samples. Moreover, administration of IPT did not influence gene expression signatures in HIV-infected participants in whom Mtb DNA was detected in PBMC at baseline, even though this intervention had previously been shown to reduce the proportion of individuals in whom this signal was detectable after treatment.^4^

The lack of association between dPCR-positivity and presence of gene expression signatures for incipient TB in our study suggests that detection of Mtb DNA in PBMC subsets of TB contacts using dPCR is more likely to represent quiescent (i.e. truly latent) Mtb infection than incipient or subclinical TB disease. This interpretation is supported by the observations that no Mtb DNA-positive participants in our study progressed to active TB over 6-month follow-up, and that prevalence of detectable Mtb DNA (79.2%) was much higher than the risk of progression to active TB (commonly estimated at ∼10%). Our findings chime with those of a linked study in which we report a lack of association between blood RNA signatures and sputum culture status in patients with active TB who have taken 8 weeks of intensive antimicrobial therapy:^15^ together, these reports provide the most direct evidence to date that Mtb can persist in humans in the absence of an active host response. They contrast, however, with results of other studies, conducted in the UK, which have reported that detection of Mtb DNA in whole blood of asymptomatic TB contacts using qPCR associates with the presence of 18F-fluorodeoxyglucose avid intrathoracic lesions on positron emission tomography–computed tomography (PET-CT) scanning, as well as increased risk of progression to active disease.^16,17^ Differing findings between these studies and our own may reflect differences in the methodology used to detect Mtb DNA in blood, since other studies^16,17^ utilised a lower blood volume than we did, and detected Mtb DNA using qPCR rather than dPCR in whole blood rather than in PBMC subsets: these factors may have reduced sensitivity, which may have contributed to the lower prevalence of Mtb DNA detection reported in asymptomatic TB contacts (16.7% in UK^16^ vs. 79.2% in Ethiopia).^4^ Contrasting findings may also reflect differences in HIV prevalence and intensity/frequency of exposure to infectious TB between participants studied in the UK vs. Ethiopia.

Our study has several strengths. We investigated a comprehensive panel of eight different RNA signatures reported to associate with incipient TB, and found a consistent lack of association with our Mtb DNA signal. Our sample size provided adequate power to detect modest differences in gene expression Z-scores between groups. Inclusion of HIV-infected and -uninfected participants, and contacts of human and animal index cases, improves generalisability of our findings. Moreover, we conducted rigorous control experiments to demonstrate that positive results from our dPCR assay are very unlikely to have arisen as a result of cross-contamination or non-specific amplification.^4^

Our study also has some limitations. Given the low-income setting, we relied on clinical symptom screening and plain chest radiographs to detect active TB: this approach is less sensitive than PET-CT scanning, especially for sub-clinical or incipient disease. However, PET-CT involves significant exposure to ionising radiation, and the lack of gene expression signature-positivity in our cohort suggests that we were successful in excluding these patients. Administration of IPT was not directly observed: given the six-month duration, it is likely that adherence was sub-optimal in at least some participants, which may have contributed to failed clearance of Mtb in some participants. The fact that we detected Mtb DNA in a significant proportion of participants completing 6 months of IPT is nevertheless consistent with results of a meta-analysis of RCTs showing that this intervention reduces risk of reactivation by only 56%.^18^

In conclusion, we report that detection of Mtb DNA in PBMC of asymptomatic adult TB contacts in Addis Ababa, Ethiopia, does not associate with the presence of host blood gene expression signatures of incipient tuberculosis, either at baseline or following completion of preventive therapy. This finding suggests that detection of Mtb DNA in PBMC subsets of TB contacts using dPCR is more likely to represent quiescent Mtb infection than incipient or subclinical disease.

## Data Availability

All data produced in the present study are available upon reasonable request to the authors

## ACKNOWLEDGEMENTS

This work was supported by a grant from the United Kingdom Medical Research Council (Reference Number MR/P024548/1, to ARM). MN acknowledges support from the Wellcome Trust (207511/Z/17/Z) and from the National Institute for Health Research Biomedical Research Funding to University College London and University College London Hospital. We thank all the people who participated in the study, and members of the field and laboratory teams who enrolled participants and processed samples.

## AUTHOR CONTRIBUTIONS

ARM, MB and STR conceived the study. MA, MB, AA, GA, MA, STR and ARM contributed to study design. JR, MB, SY, DAJ, STR, MN and ARM contributed to development of laboratory assays. MA, MB, BT, DT, MT, AA, GA and ARM contributed to implementation of the study, enrolment and follow-up of patients. MB, BT, SY, DAJ, DT, MA, STR and ARM contributed to data acquisition. JR, DAJ, MN and ARM verified underlying data of the study. JR and MN did statistical analyses. ARM wrote the first draft of the manuscript and had final responsibility for the decision to submit for publication. All authors reviewed the final draft and agree with its content and conclusions.

## DATA SHARING

A de-identified copy of the study database will be made available from the corresponding author from the date of publication.

